# A low-cost, rapidly scalable, emergency use ventilator for the COVID-19 crisis

**DOI:** 10.1101/2020.09.23.20199877

**Authors:** Samuel J. Raymond, Trevor Wesolowski, Sam Baker, Yuzhe Liu, Jordan L. Edmunds, Mauricio J. Bustamante, Brett Ley, Dwayne Free, Michel Maharbiz, Ryan Van Wert, David N. Cornfield, David B. Camarillo

## Abstract

For the past 50 years, positive pressure ventilation has been a cornerstone of treatment for respiratory failure. Consensus surrounding the epidemiology of respiratory failure has permitted a relatively good fit between the supply of ventilators and the demand. However, the current COVID-19 pandemic has increased demand for mechanical ventilators well beyond supply. Respiratory failure complicates most critically ill patients with COVID-19 and is characterized by highly heterogeneous pulmonary parenchymal involvement, profound hypoxemia and pulmonary vascular injury. The profound increase in the incidence of respiratory failure has exposed critical shortages in the supply of mechanical ventilators, and those with the necessary skills to treat. While most traditional ventilators rely on an internal compressor and mixer to moderate and control the gas mixture delivered to a patient, the current emergency climate has catalyzed alternative designs that might enable greater flexibility in terms of supply chain, manufacturing, storage and maintenance. Design considerations of these “emergency response” ventilators have generally fallen into two categories: those that rely on mechanical compression of a known volume of gas and those powered by an internal compressor to deliver time cycled pressure- or volume-limited gas to the patient. The present work introduces a low-cost, ventilator designed and built in accordance with the Emergence Use guidance provided by the US Food and Drug Administration (FDA) wherein an external gas supply feeds into the ventilator and time limited flow interruption guarantees tidal volume. The goal of this device is to allow a patient to be treated by a single ventilator platform, capable of supporting the various treatment paradigms during a potential COVID-19 related hospitalization. This is a unique aspect of this design as it attempts to become a one-device-one-visit solution to the problem. The device is designed as a single use ventilator that is sufficiently robust to treat a patient being mechanically ventilated. The overall design philosophy and its applicability in this new crisis-laden world view is first described, followed by both bench top and animal testing results used to confirm the precision, capability, safety and reliability of this low cost and novel approach to mechanical ventilation during the COVID-19 pandemic. The ventilator is shown to perform in a range of critical requirements listed in the FDA emergency regulations and can safely and effectively ventilate a porcine subject. As of August 2020, only 13 emergency ventilators have been authorized by the FDA, and this work represents the first to publish animal data using the ventilator. This proof-of-concept provides support for this cost-effective, readily mass-produced ventilator that can be used to support patients when the demand for ventilators outstrips supply in hospital settings worldwide. More details for this project can be found at https://ventilator.stanford.edu/

## 1. Introduction

COVID-19 is highly contagious and results in a wide range of respiratory distress states [1], the World Health Organization estimates that 1 in 5 adults who contract the disease will require hospitalization for breathing difficulties, and 1 in 20 will end up in the ICU under critical care, requiring a ventilator [2]. These factors mean that, as was seen during the early stages of the 2020 pandemic, and with the virus expected to persist around the world for months/years to come [3], hospitals are at risk of becoming overwhelmed with the need for both ventilators and operators who can manage patients suffering from the respiratory complications that COVID-19 can induce. If insufficient numbers of existing ventilators are present, and if there are not enough trained operators [4], patients in need of adequate ventilation will be left with a lower standard of care. The worst case for this already having been experienced in northern Italy, New York City, and areas of South America where those over 60 were essentially “left to die” [5, 6].

A low-cost, rapid to manufacture ventilator, whose functionality is able to carry a patient from early admission to their final discharge from the hospital is one such solution to this problem. This device would produce the high, continuous flow of oxygen-rich air, as used in High-flow Nasal Cannula and CPAP devices. In addition, if the patient’s condition worsened, the ventilator would also need to be able to ventilate in both assisted and mandatory modes, depending on the state of the patient. For rapid and high volume manufacturing, this ventilator would need to consist of a small number of parts, relative to existing options, and use very few custom parts.

To address the critical shortage of mechanical ventilators (MV), the FDA provided explicit details surrounding the process and requirements for devices that might eligible for Emergency Use Authorization (EUA). This Emergency Use regulatory pathway limited requirements to those vital to health and safety requirements. Key to this pathway is that the authorized products only remain in use during the Emergency. Subsequent to the emergency, products must meet all standards specific to each product and submit information to support regulatory clearance or approval from the FDA. This EUA announcement was made for many other product types in the fight against COVID-19, not just for ventilators [7]. While the existing manufacturers of ventilators were collaborating with larger corporations, such as Ford and General Motors, to utilize their massive supply chains to try and scale up production, many other smaller teams, some of whom with little or no experience in ventilators or medical equipment relied on these EUA documents to guide and optimize their designs[8]. The guiding specifications that the FDA had provided that were the most relevant for an engineering design were those listed in the Emergency Use Ventilator document [9], Table 1 shows these pertinent features. With the help of these constraints, and with the awareness that the typical supply chains would be pushed beyond capacity for ventilator-specific parts, the task then was to discern what design choices would allow for a device to be created that would fulfill the necessary constraints and still be a viable option for use in the early stages of the COVID-19 pandemic. As different teams around the country, and around the world, sought to answer this question, two distinct design paths became clear. A MV must reliably, accurately and consistently deliver a specific volume of gas in a specified time interval at specified number of times each minute. This implies that some control over the gas flow is essential. Whether the gas is first collected to a known volume and expelled (such as is the case for the “Ambu-bag” designs [10], or the gas is taken from the available sources at the time that a breath is required and valves used to interrupt the gas flow (as is the case for ours and other designs [8]), this is the fundamental design choice that separates most of the swarm of new designs that have been proposed since the beginning of this pandemic [11]. While many other designs have been produced on both of these paths, the goal for this work was to introduce a novel design that aimed to be a ‘one-device-one-visit’ respiratory care platform that is able to accompany a patient through all respiratory aspects of COVID-19 treatment. This aim goes beyond the designs that have been publicized so far as they aim to tackle the most critical-care aspect of patient ventilation, invasive ventilation. The design presented herein is capable of providing non-invasive care, such as typically provided through High Flow Nasal Canula or CPAP devices, as well as the invasive, assisted and mandatory control modes of ventilation needed when a patient no longer posseses the ability to breath on their own. This design is aimed to simplify the situations wherein a hospital is inundated with patients and the constant swapping of devices is required. This platform is also intended to lower the barrier for respiratory care during COVID-19 treatment to enable a larger workforce of medical professionals to participate to patient care. The document is presented as follows: the design of this ventilator is described including the design philosophy used to overcome the unique constraints in an EUA/COVID-19 situation, advantages and disadvantages of this approach are discussed. Next the device is shown to operate over the conditions expected during a full hospitalization using bench-top equipment to measure the relevant parameters from the ventilator. Finally a live animal experiment is presented and the results analyzed to determine the potential dangers of ventilation using this device to detect any causes of ventilator-induced lung injury, using a porcine subject. Finally, a discussion of the use of this ventilator platform and any relevant conclusions are presented.

**Table 1:** Emergency Use Ventilator, minimum requirements for treating COVID-19 patients [9].

| EUA Property | Recommended Value |
| --- | --- |
| FiO<sub>2</sub> (fraction of inspired Oxygen) | 21% (ambient) to 95% of the source oxygen concentration input no more than 10% steps |
| PEEP | 5 to 20 cmH<sub>2</sub>O in no more than 5 cmH<sub>2</sub>O steps |
| I:E Ratio | 1:2 preferably adjustable from 1:1 to 1:3 |
| Mandatory Ventilation Mode | respiratory rate from 10 to 30 inflations/min steps of no more than 2 inflations/min |
| Tidal Volume | 350 to 450 ml +/- 10% in no more than steps of 50 ml, preferably a lower range of 250 ml and an upper range of 600 ml or 800 ml |
| Inspiratory Pressure Limit | 15 to 40 cmH<sub>2</sub>O steps of no more than 5 cmH<sub>2</sub>O |
| Alarm Conditions | · Gas or electricity supply failure.<br>· Ventilator switched off while in mandatory ventilation mode.<br>· Inspiratory airway pressure exceeded.<br>· Inspiratory and PEEP pressure not achieved (equivalent to disconnection alarm condition).<br>· Tidal volume not achieved or exceeded. |
| Visual Indicators | The current settings (e.g., inspiratory pressure, tidal volume, frequency, PEEP, FiO<sub>2</sub>, ventilation mode).<br>The current delivery (e.g., inspiratory pressure, tidal volume, respiratory rate, PEEP, and FiO<sub>2</sub> at the patient-connection port). |

## 2. Methods

With the circumstances surrounding the emergency/rapid-response ventilator innovation period that occurred in the Spring of 2020, certain design conditions were put in place both by the FDA regulations and by the nature of the emergency itself. A complete but scalable design was required to reach as many people as possible. Many different designs have been authorized by the FDA for the EUA and many will likely not be utilized. Our design is intended to exist after the EUA phase has passed and as such this design required some extra thought to its usefulness in the long term.

### 2.1. O2U Ventilator Design Philosophy

The O2U ventilator is based on a continuous flow design, where the valves direct flow into or away from the patient’s breathing circuit during invasive ventilation modes, and also allow for continuous flow during non-invasive mode operation. This is achieved by connecting the ventilator to pressurized sources of Air and Oxygen gas. These gasses are pre-mixed to the desired fractional concentration of Oxygen (FiO_2_) before entering the ventilator. During non-invasive operation modes this gas mixture is allowed to pass freely through the device, entering and exiting the patient breathing circuit, operating in the same manner as other CPAP or High Flow Nasal Canula systems. Inspiratory and expiratory valves are kept open in this state and the system monitors the pressures in case of leaks or blockages or any other risks to patient and device safety. For invasive ventilation modes, either assisted or mandatory, the inspiratory and expiratory valves are used to allow or prevent the passage of incoming and outgoing gas, respectively, to enforce the respiration cycle within the patient. To overcome the lack of availability in flow-measuring sensors, this ventilator relies on a Pressure-Limited-Time-Cycled breathing loop where the flow rate is set on the ventilator using a manual control valve, also known as a Thorpe Tube, and the inspiratory time is set such that a known volume of gas is delivered during each inspiration phase. A specific tidal volume can then be delivered by knowing the flow rate and adjusting the inspiratory time such that the tidal volume (V_*T*_) can be calculated simply as *V*_*T*_ = Flow rate *×* Inspiratory Time. where the operator can determine, for a user set flow rate and inspiratory time, the delivered flow volume. Volume calculations are still important for patient care, so in addition to this table for setting the desired delivered volume, a spirometer-based expiratory volume sensor is used on the expiration side of the circuit to measure exhaled volumes. This spirometer-based volume sensor is based on the work of Edmunds et.al [12], who’s work was aimed to help measure gas volumes in ventilators for the COVID-19 pandemic. The O2U ventilator prototype is shown in Figure 1.

**Figure 1:**
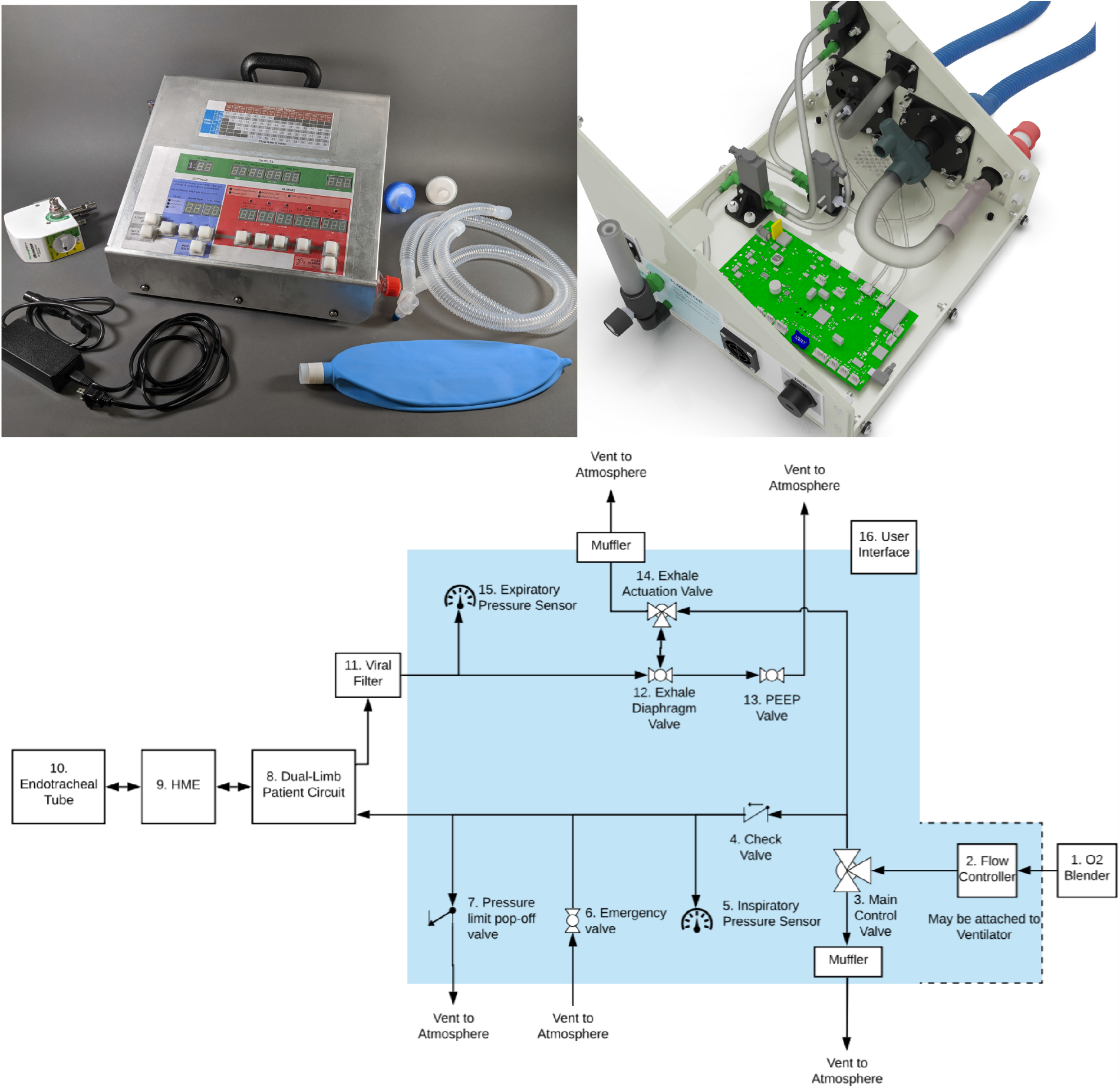
A prototype of the new O2U ventilator, designed and built in the early months of the COVID-19 pandemic with accessories(top-left) an internal construction render (top-right) and the prototype schematic (bottom) showing the internal components (within the shaded region) and the relevant accessories and components required (non shaded).

### 2.2 Advantages and Disadvantages

With any of the proposed ventilator designs that were published to meet this challenge, each of these approaches required compromising on some feature or price point in order to meet the best possible match of price to performance, however in addition to this, other constraints such as supply chain and clinical need meant that other choices were required that further added to the constraints and limitations for each design. The O2U ventilator design consists of a number of unique choices that were done to maximize the utility of the ventilator to act as a one-device-one-visit platform. Below are two of the design choices that led to the most impactful differences between this design and the majority of other proposed ventilators.

#### 2.2.1 Reliance on externally pressurized and mixed gas

One potential limitation of the O2U ventilator is the reliance it has on a pre-mixed and pressurized input of gas into the system. With the assumption being that most hospitals and care facilities will have access to both an Oxygen/Air blender and pressurized gasses, the benefit is that the ventilator can be made with far fewer parts and can utilize the pressure of incoming gas to pressurize the circuit. The input Thorpe tube regulates the pressure from wall pressures of typically 50psi down to 1-3psi, the regular range for patient breathing circuits.

#### 2.2.2 Pressure-Limited-Time-Cycled ventilation

Most ventilators offer pressure-controlled or volume-controlled ventilation. This requires the use of a closed feedback look in the case of pressure-control, or a flow sensor in the volume-controlled case. The O2U ventilator operates in a modified manner to these two common cases and instead relies on a known flowrate entering the system and control of the valve timings. This control type: Pressure-Limited-Time-Cycled ensures that the pressures are always monitored to detect any leaks or obstructions that could risk patient or device safety, and that the desired delivered volume is instead controlled by the amount of time that the inspiration valve is open for. This assumption that the gas will be a constant input allows this ventilator to be used in non-invasive mode and invasive mode. The non-invasive case requires a mask be placed over the patient’s mouth and the continuous flow is channeled through the mask and allowed to exit the expiratory valve, enabling the patient to take a breath of enriched/higher pressure gas that will lower the breathing effort. For the invasive ventilation where a patient is intubated the timing of the valves directs the flow into or away from the patient. A tidal volume is set by allowing a known flowrate of gas to flow for a specified time.

### 2.3 A single ventilation platform to manage COVID-19 patients

A typical patient who would suffer severe COVID-19 symptoms would follow a trajectory similar to that described as follows. A patient arrives at a hospital feeling weak and suffering from impaired breathing, but able to breathe on their own. They would normally present with low Oxygen saturation and be placed on a nasal canula/ CPAP or BiPAP machine with a higher percentage of Oxygen, potentially up to 100%. After some time the patient may continue to decline and become weaker and confused as the toll on their body leads to an increased work of breathing. The patient’s lungs and airways have become compromised with mucus build up and general weakness, so the patient is placed under anesthesia and intubated to allow for a mechanical ventilator to be connected to assist in breathing, a variety of assisted and mandatory modes may be used to properly manage the patient, at the discretion of the care provider [13]. After such time as their body has been able to fight off the disease and the ventilator has protected their lungs as best as possible, weaning off the ventilator will occur and the patient will begin to take more spontaneous breaths. Eventually, before the patient is able to be extubated completely, they may be placed on a tracheal collar for passive Oxygen (still invasive). After some more time to build strength such that they could safely undergo extubation, they would likely require more non-invasive assistance for breathing, such as the High Flow Nasal Canula or CPAP/BiPAP devices used at the beginning of their hospitalization. Finally, the patient would recover sufficiently to breath completely unassisted and subsequently be discharged. While this is not the case for every severity level of COVID-19 patients, this represents one such path that highlights the number of different breathing devices that are possible when being treated for this disease.

The goal of the O2U ventilator is to simplify this process and enable a single device to follow along with the patient as they progress with their treatment. This minimizes the number of different connections and devices that a medical professional needs to work with and maximizes the familiarity that the operator can have with the device which is important for those with less experience treating critically ill patients with ventilators. To show the abilities of this ventilator to manage these tasks a number of bench-top tests and animal testing was conducted to observe the flexibility of this ventilator in the cases where it was designed to be used. The animal test in particular was used to highlight the performance of the ventilator and any potential risk of damaging normal lungs.

### 2.4 Bench Top Testing - Protocol

To ensure that the O2U ventilator met the EUA requirements needed to treat COVID-19 patients (any other aspects of FDA/regulatory compliance will not be discussed here) a number of tests were performed on the ventilator and the data recorded to ensure that these requirements could be met. Specifically, the following variables relevant to COVID-19 treatment were tested:

- Positive Expiratory End Pressure (PEEP): to ensure that the lung always has some positive pressure inside so that the lung, which can become filled with fluid does not collapse on itself during exhalation, a small amount of PEEP is often used to protect the lungs.
- Breathing Rate (breaths/min delivered): depending on the vitals of the patient, their lung characteristics, and the current state of the ventilator settings, different breathing rates (how often the ventilator pushes air and Oxygen into the lung) are required.
- Tidal Volume: Lung volumes typically relate to overall body size and hence the tidal volume (the total volume that the ventilator sends during inspiration) needs to be variable to treat patients of different ages, genders, and sizes.
- Inspiratory Time (similar to I:E ratio): based on the breathing rate, a fixed amount of time is placed between breaths, this can be split into inspiration and exhalation phases at a ratio, normally more time is given for exhalation as the lungs contract slowly compared to the ventilator valves.

Tests were conducted using a Michigan Instruments test lung and two TSI flow measuring devices. The protocols for these parameter sweeps are outlined in Table 2. Results for the performance of the ventilator are shown in the next section.

**Table 2:**
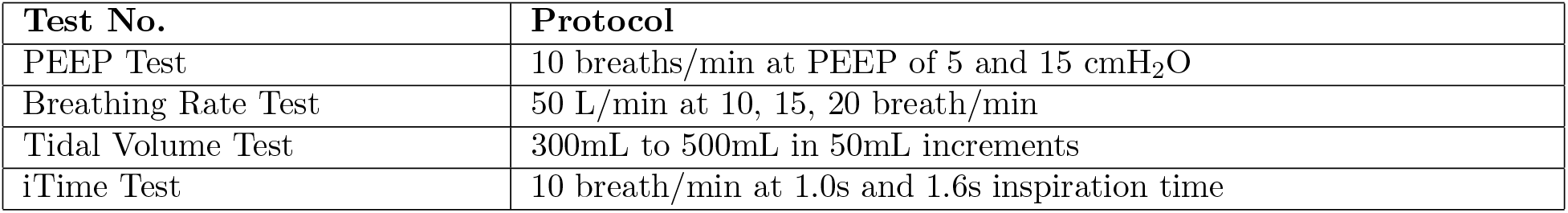
Bench-top tests for different parameter values for the O2U ventilator.

### 2.5 Live Animal Testing - Protocol

During ventilation, two types of injuries may be induced by mechanical ventilation: (a) high inspiration lung volume or pressure leading to alveolar overdistension, in which the lung tissue is damaged; (b) cyclic change in the nonaerated lung, for which the underlying cellular mechanism is still unclear. To prevent injury (a), tidal volume should be smaller than a critical value to avoid large stresses in the lung tissue. To prevent injury (b), PEEP should be taken to avoid low end-expiratory lung volume (EELV), which will lead to high strains in the lung. The upper limit of the tidal volume will also help to avoid high strains, therefore it also helps to prevent the injury (b). To test this ventilator design and to add additional information beyond the minimum required by the EUA, a porcine animal study was conducted where a pig was ventilated under a number of conditions by the O2U ventilator in order to judge the potential for lung damage in the ventilator’s operation. Pigs have been proved as an effective large animal model for ventilation in previous work [14]. Furthermore, humans and pigs have similar respiratory rates, which is an important parameter for replicating similar air circulation during breathing. The protocol for the animal test is shown in Table 3.This protocol was designed to both test the usability of this ventilator in a real-world scenario with a live subject, and to take the animal through various phases, representative of the cases that can occur in typical respiratory care where larger and smaller tidal volumes are prescribed during treatment. The animal was a 65kg female pig with normal lungs. Arterial and venous blood lines were drawn from the animal periodically to monitor the effect of the ventilation during each phase of the experiment. A baseline of 10ml/kg of tidal volume, as reported in [15] was used as a baseline for this test with the hyper- and hypoventilation deviations based on 30% increase and reductions from this amount, respectively. Additionally, for this test, the animal’s lungs were extracted after the experiment to observe any gross signs of injury. Results of this animal test are shown in the next section.

**Table 3:** Protocol used during the O2U animal test.

| Phases for Animal Test | Protocol |
| --- | --- |
| Baseline | Ventilator Setting: 10bpm, 1.2insp, 33L/min, PEEP 5cmH<sub>2</sub>O |
| Hyperventilation | Ventilator Setting: 10bpm, 1.6insp, 33L/min, PEEP 5cmH<sub>2</sub>O |
| Hypoventilation | Ventilator Setting: 10bpm, 0.8insp, 33L/min, PEEP 5cmH<sub>2</sub>O |
| Baseline/Recovery | Ventilator Setting: 10bpm, 1.2insp, 33L/min, PEEP 5cmH<sub>2</sub>O |

## 3 Results

### 3.1 Bench Top Testing - Results

Data was recorded on the TSI flow meters and processed as comma separated value text files. These files were then post-processed using custom Python scripts to analyze the flow data. Flow of gasses were recorded, along with the pressure. These pressures were measured as absolute values and atmospheric pressure was measured during testing to calculate the gauge pressure, measured in cmH_2_O. All of the tests listed in Table 2 and the analyzed data is shown in Figs 2 - 5 for PEEP, breathing rate, tidal volume, and inspiratory time, respectively. Data was recorded over several breaths for each test, representative plots are shown here for clarity.

**Figure 2:**
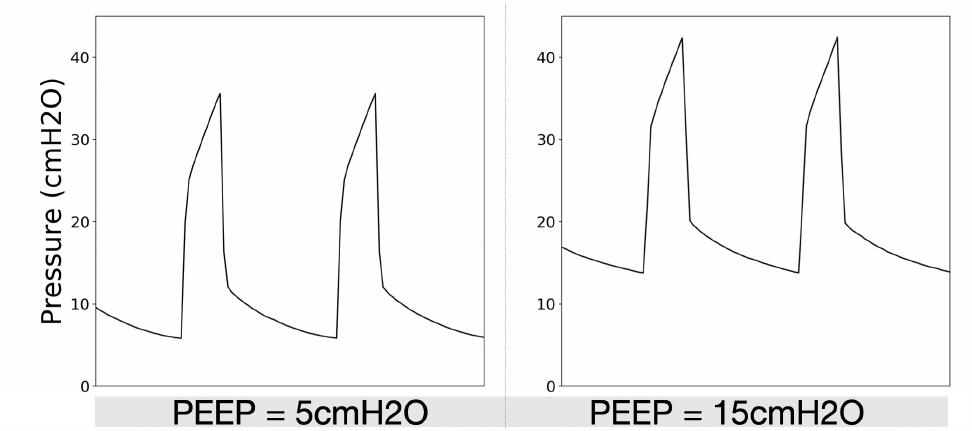
Positive end-expiratory pressure (PEEP) as a function of time as measured on the bench-test setup for the O2U ventilator. (left) PEEP at a level of 5cmH2O and (right) PEEP at 15cmH2O set via an external diaphragm valve on the ventilator.

**Figure 3:**
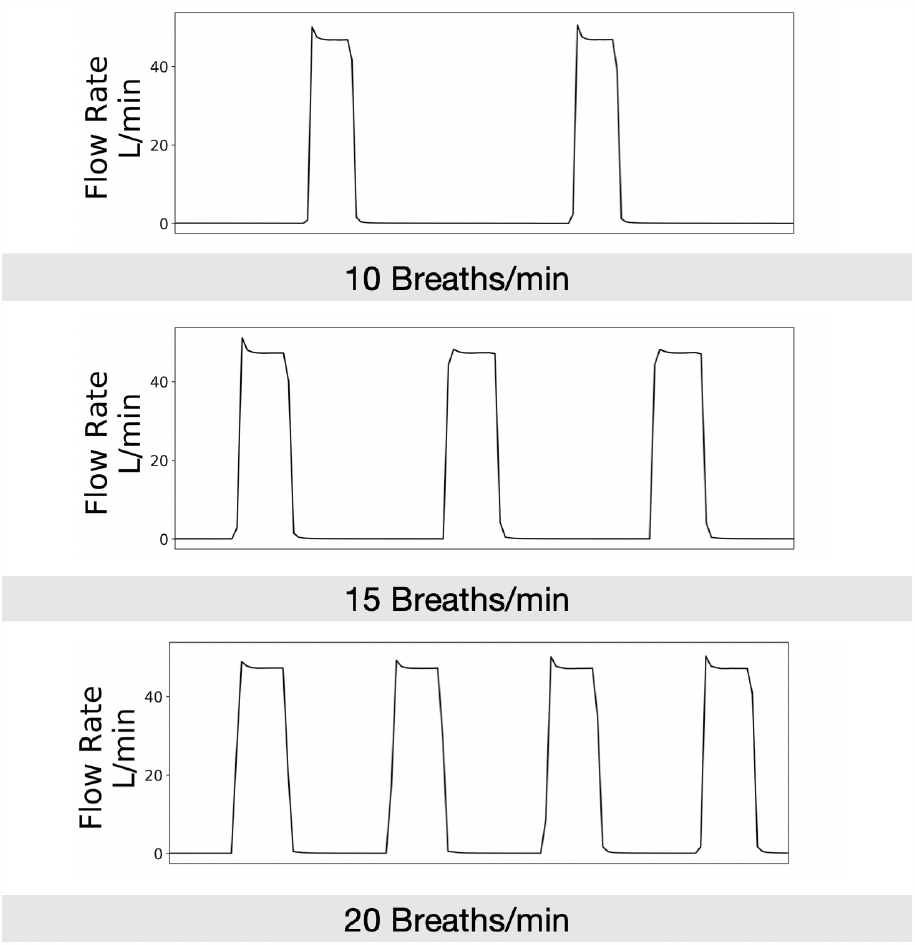
Breathing rates control how rapidly air is supplied to the patient. Control of the breathing rate is essential for patient case. Three graphs showing 10 (top), 15 (mid), and 20 (bot) breaths per minute as set by the UI of the O2U ventilator for a flowrate of 50L/min.

**Figure 4:**
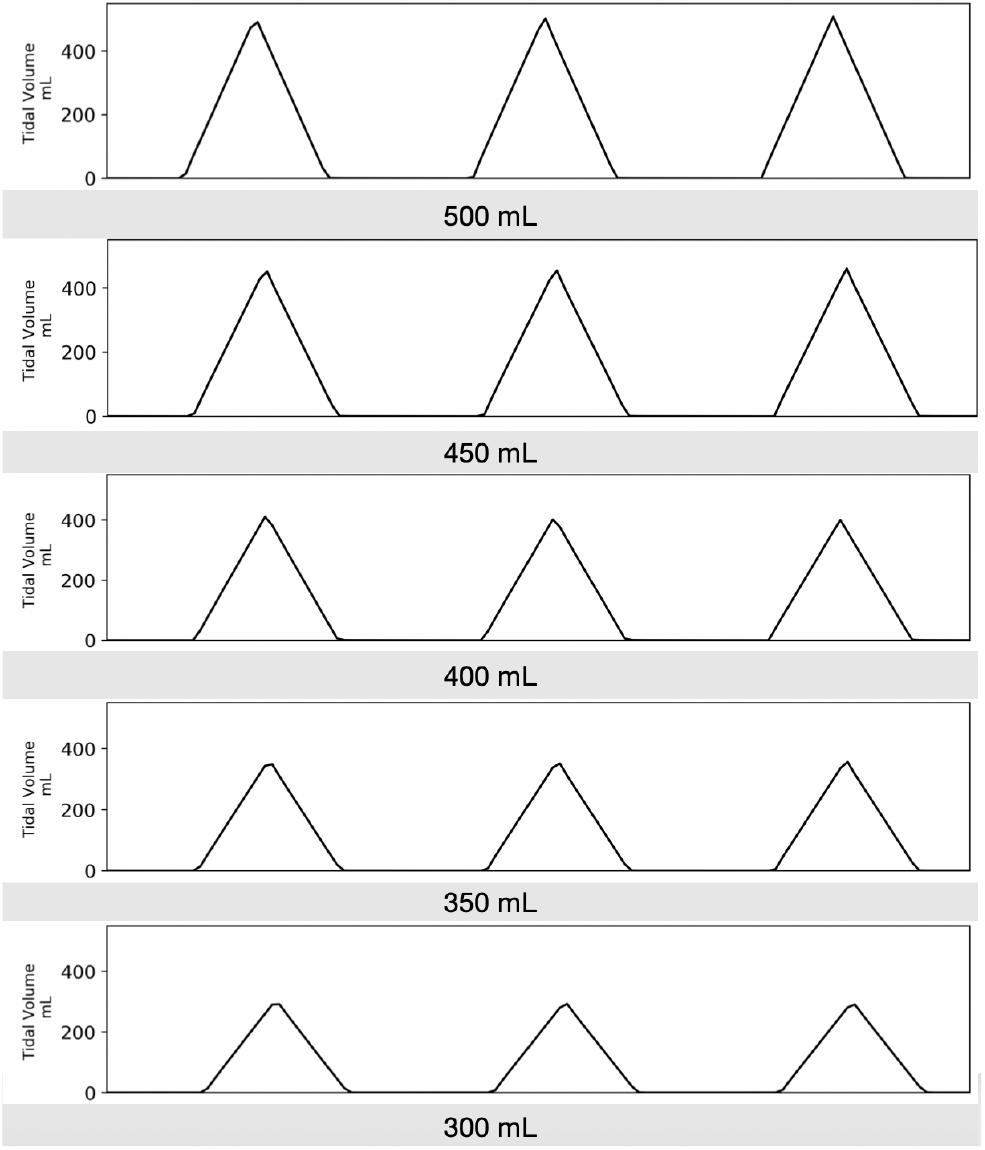
Tidal volume, or the volume delivered to the patient is a critical variable in patient care. The O2U ventilator is capable of providing the FDA-required ranges to manage the majority of COVID-19 patients. This range is shown for 50mL increments from 300mL to 500mL.

**Figure 5:**
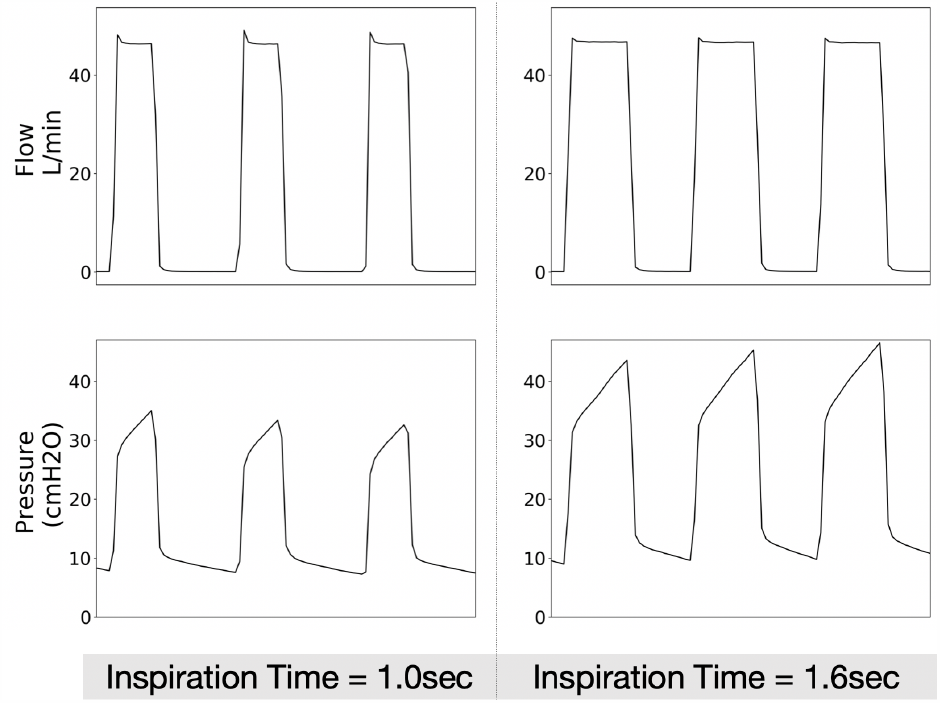
Inspiration time is the duration of time that the air is delivered to the patient. This can be important to vary to manage different lung care strategies. Shown here is the difference in 1.0s and 1.6s on the dynamic effects on the flow and PEEP pressure signals.

**Figure 6:**
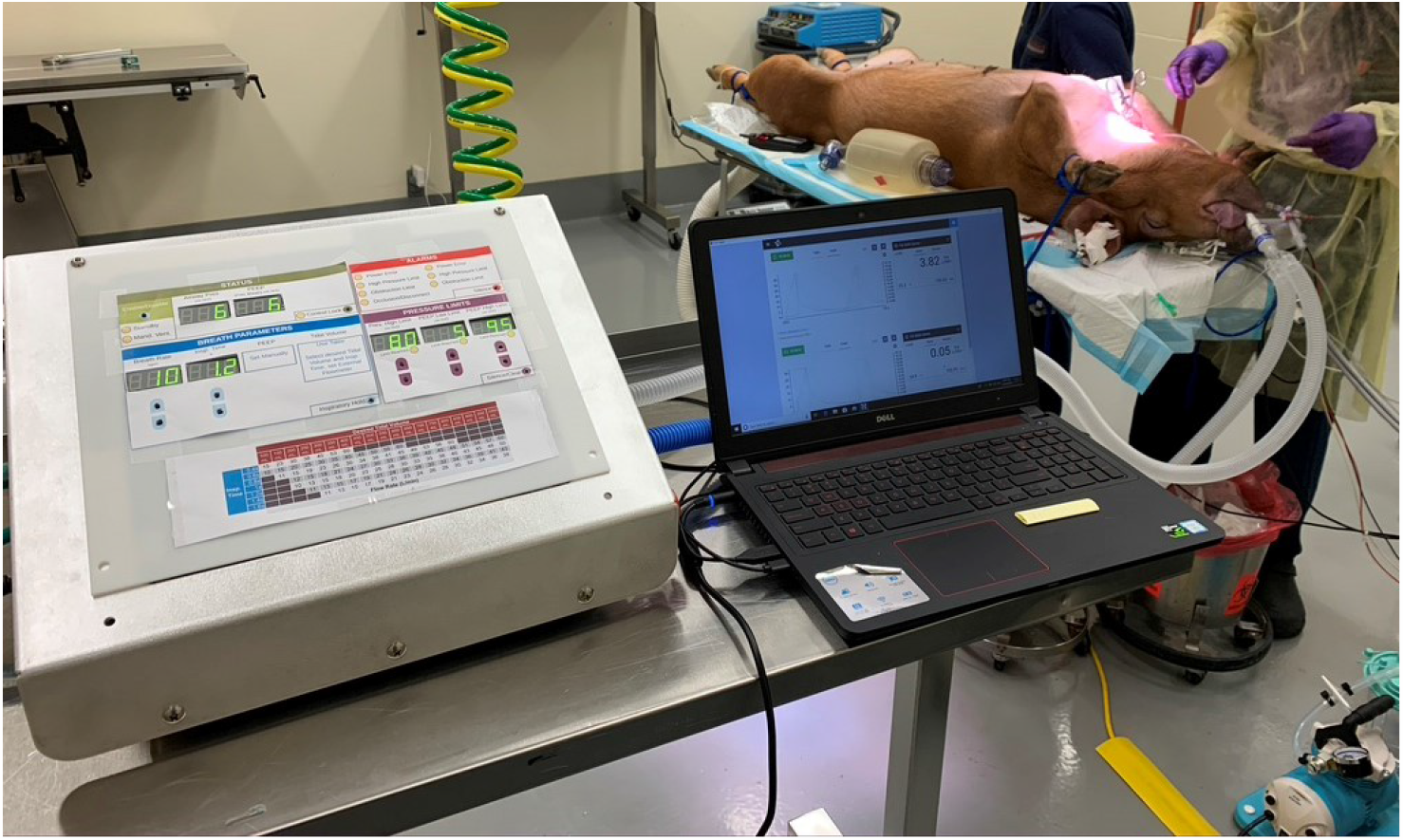
The O2U ventilator performing mandatory ventilation on a sedated and intubated porcine subject.

### 3.2 Live Animal Testing - Results

For the animal test, the subject was sedated and intubated and immediately placed on the ventilator at the baseline rate of 10 breaths/min and 650mL of tidal volume. Blood gasses were taken every 15 mins, with the arterial data shown in Table 4 and the venous gasses shown in Table 5.

**Table 4:**
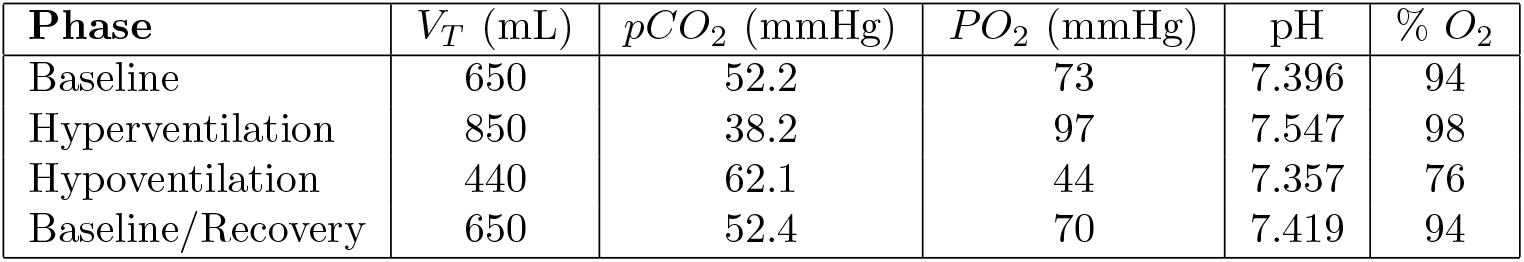
Arterial blood measurements during the pig experiment.

**Table 5:** Venous blood measurements during the pig experiment.

| Phase | $V_T$ (mL) | $pCO_2$ (mmHg) | $PO_2$ (mmHg) | pH | % $O_2$ |
| --- | --- | --- | --- | --- | --- |
| Hyperventilation | 850 | 43.4 | 48 | 7.488 | 86 |
| Hypoventilation | 440 | 66.2 | 38 | 7.330 | 66 |
| Baseline/Recovery | 650 | 56.5 | 46 | 7.400 | 80 |

Data on the flow and pressure on the inspiratory side was measured using the same TSI flow meter used for the bench-top testing. Flow, pressure, and delivered volume for each phase listed in Table 3 are shown in Figs 7 - 9, respectively.

**Figure 7:**
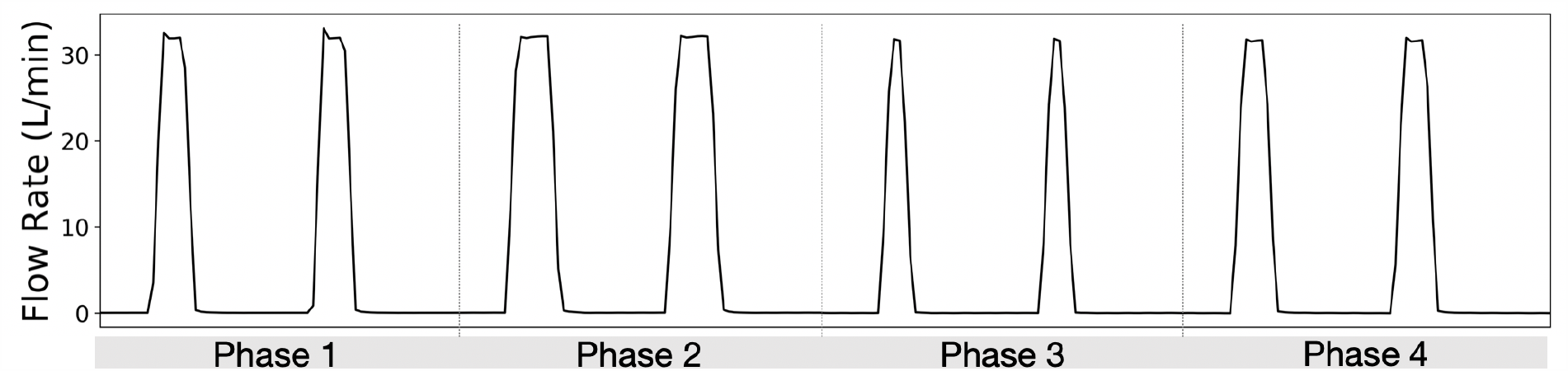
During the animal test, while the flowrate was kept constant at 33L/min, due to the changes in other operating parameters, the flow signals were different for each phase of the test.

**Figure 8:**
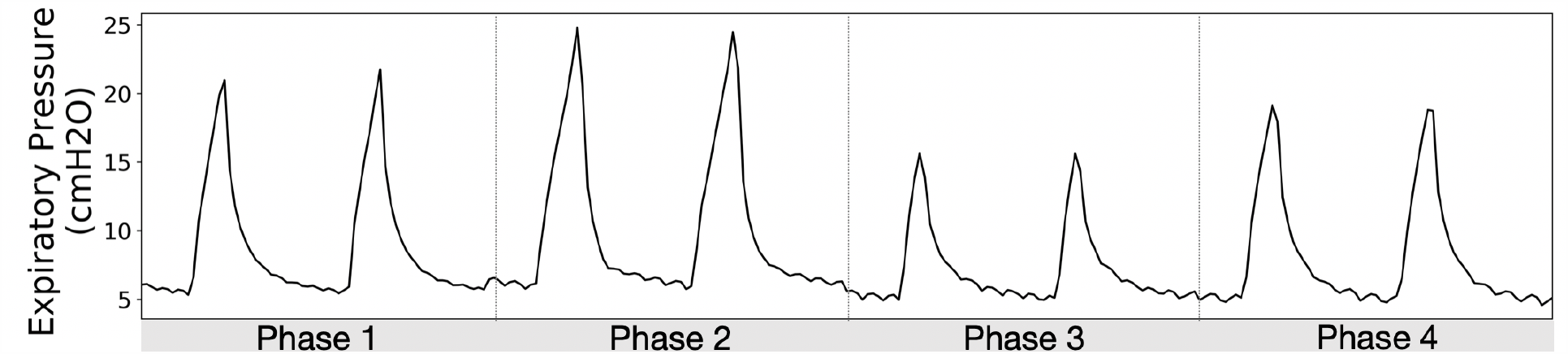
While the PEEP pressure was maintained at 5cmH2O for the animal test, during each phase the pressure peaks and duration changed in accordance with the ventilator setting and the animal response.

**Figure 9:**
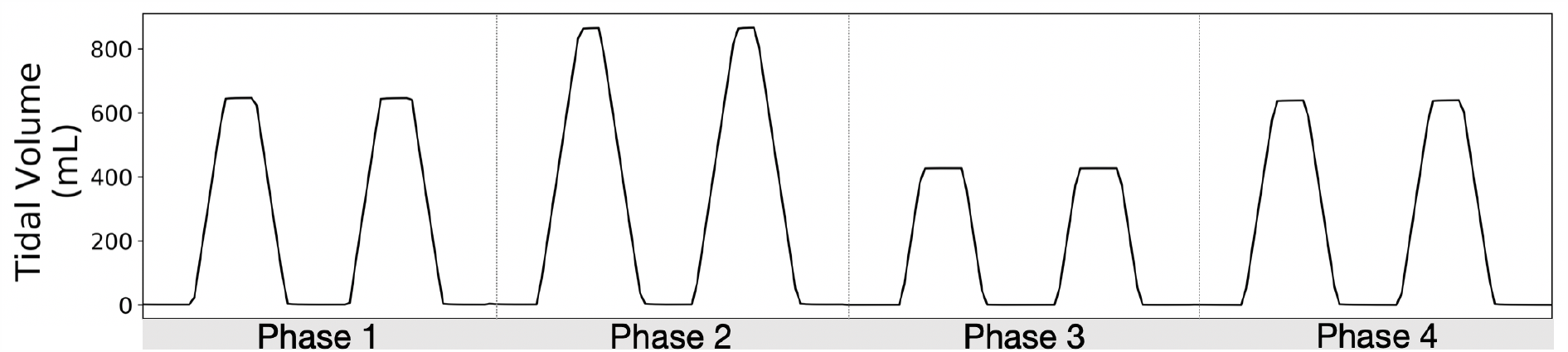
Delivered/Tidal volumes were the key variable in the animal test, here each phase consists of a different volume to induce the measured responses in the animal that are shown in Table 4 and 5.

### 3.3 Lung Histology

In addition to the blood gasses measured during the experiment, after the ventilation, the animal was euthanized and X-Rays and gross histology of the lungs were performed to ascertain the extent of potential damage caused by the ventilator. The X-Rays, shown in Figure 10 and the histology report of the lungs indicate that there was some reddening/congestion in the caudal dorsal lung fields (see Figure 11), which may have been positional as the pig was in dorsal recumbency during the procedure. There was no evidence of hyperinflation or atelectasis. These results indicate that the lungs were well preserved by the ventilator during the experiment.

**Figure 10:**
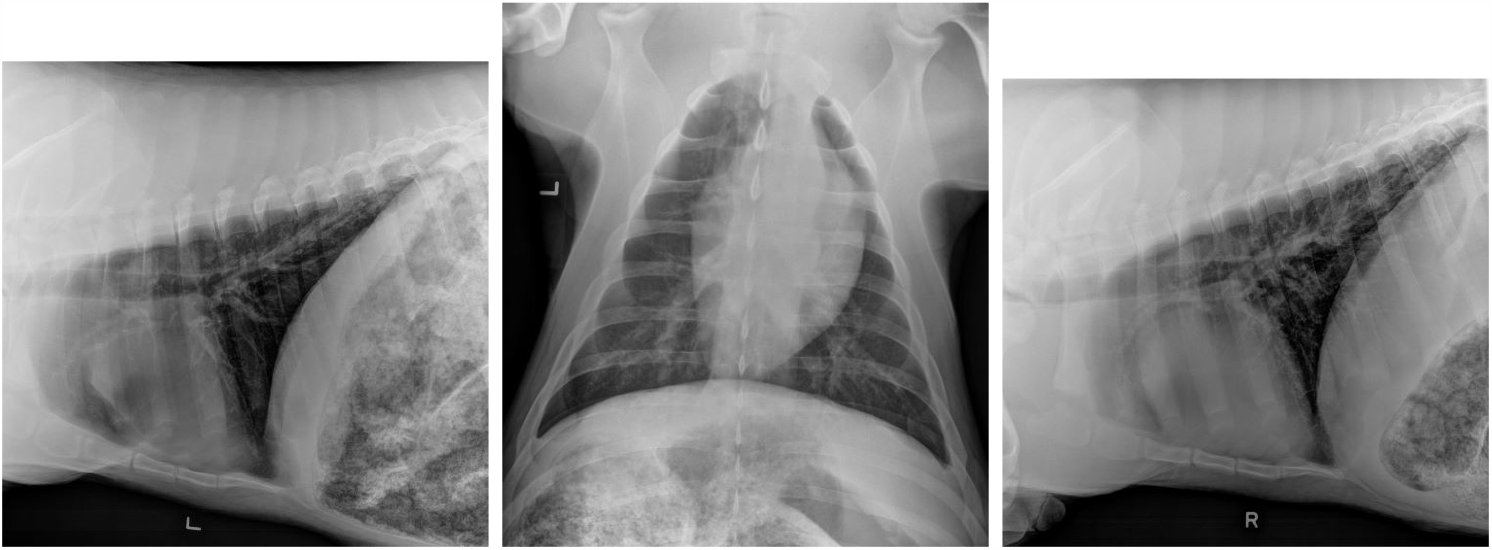
X-Ray images of the animal lungs after the ventilator experiment. (left) Left Lateral Thorax, (middle) VD thorax, (right) Right Lateral Thorax.

**Figure 11:**
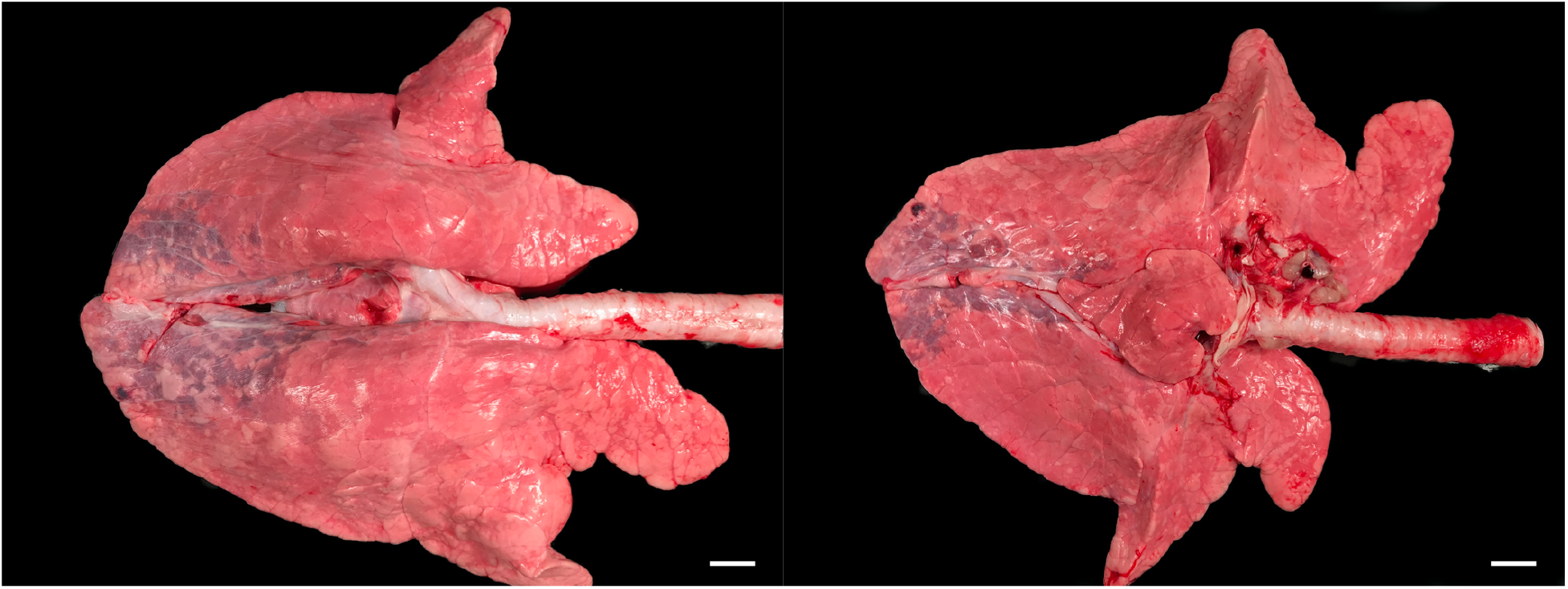
Images of the animal’s lungs after the experiment. Scale bars are 2cm.

## 4. Discussion

The results from the previous section show the ability of the O2U ventilator to perform the necessary functions, as per the FDA and reveal it’s efficacy to ventilate a living subject throughout a variety of respiratory states. The FDA’s EUA guidance provided valuable details on the scope of features and the ranges of parameters that a viable, COVID-19 ventilator requires. The ranges listed in Table 1 and the subsequent plots of the data collected in Figures 2 to 4 demonstrate the ventilator’s response to the required input as stated by the EUA guidelines. While the guidelines posted by the FDA represents the minimum requirements to pursue emergency use authorization, the O2U ventilator was also tested in a real-world setting with a living subject under mandatory ventilation. During the procedure, the animal was forced into both hyperoxic ans hypoxic states, as can be seen in phases 2 and 3 from Table 4, and the first two phases of Table 5 from both the arterial and venous blood gasses, respectively. The pH levels of the blood rose in accordance with the increase in Oxygen supplied via the larger tidal volumes, which can be seen in the plots in Figure 7. The larger inspiration time allowed the tidal volume to increase approximately 30%, resulting in the decrease of pCO_2_ during the hyperventilation phase from a baseline of 52.2mmHg to a low of 38.2mmHg, a decrease of roughly 27%, which is in line with the proportional response of the increased tidal volume. Additionally, during the hypoventilation phase, where tidal volume was decreased from the baseline of 650mL to 440mL, the pCO_2_ peaked at 62.1mmHg, an increase of 19%, also in line with the expected proportional response to this decrease in tidal volume. During the final phase of the procedure, the animal was brought back to the baseline settings to observe the recovery of the state of the animal. Both the arterial and venous measurements of the blood gasses showed a stabilization of the animal to it’s baseline state with a final pH of 7.4 for both arterial and venous blood gasses and a pCO_2_ above 50mmHg for both arterial and venous gasses. This is important as the venous line indicates that the heart was also able to recover to the baseline state during this recovery phase. The ventilator was able to provide consistent and reliable delivery of oxygen to the subject during the procedure, allowing the recovery from this wide variation in state in a relatively short amount of time. Additionally, after the procedure and the subject was terminated, the post mortem study of the lungs revealed no obvious signs of injury, despite the wide variation of ventilator settings and extreme physiological states that the animal underwent. This is a promising sign for the O2U ventilator as a rapid response tool in the COVID-19 crisis.

With the second waves of the virus spreading across the globe and with the winter months approaching for much of the world’s population, it is likely that a renewed spike in the need for MV will come at the end of 2020. New ventilator designs, such as ours shown herein will be vital to meet the need that has skyrocketed during this pandemic. Importantly, though, the designs that have spawned in the wake of this crisis will need to be properly vetted to ensure that resources are not wasted and that the health care professionals are given the right tools to manage the situation. Special attention will need to be given to which designs, even after they are awarded EUA, are taken to market for mass production and distribution. Those with sufficient evidence to show that the design has been field tested to some degree, such as the results shown in this work, represent a necessary minimum to provide some peace of mind for future users. As with the now dozens of new designs that have emerged in the last 6 months to combat this crisis, the next stage of ventilator uptake will require an investigation of appropriate use and logistics of acquisition. It is likely that the majority of the designs will not see widespread use, either in the US or abroad. However, this crisis has shown that the ventilator market, in general, was prime for disruption, and we are only beginning to see where this new paradigm will lead designers, engineers, and medical professionals as the COVID-19 crisis continues.

## 5. Conclusion

This paper introduces a low-cost, versatile ventilator designed and built in accordance with the Emergence Use guidance provided by the US Food and Drug Administration (FDA) wherein an external gas supply supplies the ventilator and time limited flow interruption guarantees tidal volume. The goal of this device is to allow a patient to be treated by a single ventilator platform, capable of supporting the various treatment paradigms during a potential COVID-19 related hospitalization. This is a unique aspect of this design as it attempts to become a one-device-one-visit solution to the problem, whereas other published, rapid response devices have focused only on the most critical use cases. The design philosophy of the ventilator is based on a continuous flow, Pressure Limited Time Controlled format and the parameters of the device are tested in accordance with the FDA’s Emergency Use Authorization guidance. Additionally, to test the device on a living subject, a pig is sedated and placed on mandatory ventilation while the ventilator controls are adjusted to bring the animal to both hypoxic and hyperoxic states until it is brought back to a healthy baseline. This test is used to detect risk of lung injury and after a post mortem, the lungs were found to be well protected by the ventilator during this procedure. The ventilator is shown to be a promising candidate for emergency use during the COVID-19 pandemic and beyond in cases where a rapid-response and versatile ventilation platform are needed.

## Data Availability

N/A

## Acknowledgments

This project was borne out of an extraordinary time and would not have succeeded without the generous support of the ChanZuckerberg BioHub, Stanford University, as well as everyone who put in time, effort, and much needed wisdom: Larry Miller (219 Design), Dave Bim-Merle (219 Design), Alex Wood (219 Design), Wes Waugh (219 Design), Andrew Sauter (219 Design), Abe McKay (219 Design), David Gutierrez (219 Design), Peggy McLaughlin (MPMAdvisors), Mike Horzewski (O2U Inc.), Christoph Mack (Andrews-Cooper), Anessa Chilton (Andrews-Cooper), Tom Ryan (Andrews-Cooper), Tyler Smith (Andrews-Cooper), Chelsea Ramm (Product Realization Group), Aadel Al-Jadda, Michael Keer (Product Realization Group), Matthew Callaghan (OneBreath), James Kennedy Wall (Stanford Children’s Health), Enrique Romo, Michael Fanton (Stanford Bioengineering), Gus Domel (Stanford Bioengineering), Hossein Alizadeh (Stanford Bioengineering), Sam Monga, Nicole Batista (MCRA), Michael John (MCRA), Bryan Loomas, Lina Miyakawa (Mount Sinai), Ed Carryer (Stanford Mech. Eng.), Sam Baker (Stanford Comparative Medicine), Godfrey Mungal (SCU Engineering), Ross Venook (Stanford Bioengineering),Jim Chua (Retired RT), Steve Quake (Stanford Bioengineering), Gajus Worthington (CZBioHub), Gregory Burns (UCSF), Polina Murdakhayev (UCSF), Brian Daniel (UCSF), Chris Danek, Scott Murano (Wilson Sonsini), Dominique Filloux, Shilpan Amin (General Motors), David Piech (UC Berkeley/UCSF Bioengineering), Jonathan Schor (UCSF/MSTP), Ryan Herbst (SLAC), Aaron Roodman (SLAC), Jon Saltonstall (AlvaMed), Mark Cox (AlvaMed).

